# Unique monocyte transcriptomic profiles are associated with preclinical atherosclerosis in women with systemic lupus erythematosus (SLE)

**DOI:** 10.1101/2020.08.05.20169136

**Authors:** Laurel Woodridge, Elvira C Chocano, Paul Ashford, George Robinson, Kirsty Waddington, Anisur Rahman, Christine Orengo, Elizabeth C Jury, Inés Pineda Torra

## Abstract

Women with Systemic Lupus Erythematosus (SLE) show significantly increased cardiovascular risk compared to the general population. However, despite CVD being a major cause of morbidity and mortality for these women, this increased risk is not managed clinically and tools to dissect and predict their cardiovascular risk are lacking. Notably, this elevated CVD risk is not captured by traditional risk factors. To explore molecular programs underlying asymptomatic atherosclerosis in SLE we used a well-characterised cohort of CVD-free women with SLE, scanned for asymptomatic atherosclerotic plaques using non-invasive ultrasound imaging of the carotid and femoral arteries. We investigated the transcriptomic profiles of CD14+ circulating monocytes in women with SLE with or without preclinical atherosclerosis. We identified unique monocytic gene expression profiles that distinguished the presence of preclinical plaques in women with SLE. In addition, advanced bioinformatic analysis revealed functional pathways and interactions between the genes identified that could explain mechanistic differences in plaque formation. We propose that these molecular signatures could help understand why a subset of women with SLE are predisposed to develop atherosclerosis and at higher risk of developing clinical CVD. Collectively with other efforts, these molecular insights will help to better define atherosclerosis in the context of SLE which will be critical for future patient stratification and identification of anti-atherosclerotic therapies.

## Introduction

Systemic lupus erythematosus (SLE) is a chronic, inflammatory auto-immune disease disproportionately affecting women (incidence of 9:1, women-to-men ratio). An unpredictable course of flares and remissions and varied symptom severity reflect the heterogeneous nature of this disease. SLE pathogenesis is attributed to the production of antibodies with specificity to a range of nuclear self-antigens and activation of effector T lymphocytes, which form immune complexes that are deposited into multiple organs, further activating immune responses (Arbuckle et al., 2003; Bayry, 2016). The progressive loss of tolerance to nuclear antigens and the accumulation of immune complexes due to impaired clearance mechanisms are central features of the disease and contribute to greater clinical severity.

A major additional risk factor in SLE and a leading cause of mortality is cardiovascular disease (CVD) and accelerated atherosclerosis, with asymptomatic atherosclerotic plaques presenting prior to CVD onset (Björnådal et al., 2004; Schoenfeld et al., 2013). Atherosclerosis results from a complex process involving abnormal or dysregulated circulating lipids and exacerbated inflammation, leading to the uptake of modified lipoproteins and accumulation of immune cells within the arterial wall (Moore et al., 2013). Specifically, an early stage in this process is the recruitment of circulating monocytes to the subendothelial space, subsequently differentiating into macrophages. Due to dysregulated cholesterol efflux these cells become lipid-laden or “foamy”, showing impaired migratory capacity and enhanced secretion of pro-inflammatory cytokines which further perpetuate an inflammatory milieu. Apoptosis and necrosis of lipid-saturated macrophages form a necrotic core which may become unstable and prone to rupture, potentially causing strokes or myocardial infarction (Moore et al., 2013). Despite a prevalence of classical CVD risk factors in SLE patients, this alone does not account for the ~50% increase in CVD-risk observed (Schoenfeld et al., 2012). This is particularly evident in the striking increase in female susceptibility in this population, with a 4-fold increase in myocardial infarctions (Esdaile et al., 2001). Despite this evidence, a female-focused mechanistic model to define CVD-risk in women with autoimmunity is lacking. Traditionally, CVD has been considered to predominantly affect men, and consequently, women have been understudied, underdiagnosed and undertreated (Balder et al., 2015; Wenger, 2018). Remarkably, over the past 10 years women have been underrepresented in trials supporting FDA-approval of new cardiometabolic drugs (Khan et al., 2020).

Likely because of the lack of appropriate stratification tools to predict CVD in women with SLE, treatment with drug therapies targeting lipid metabolism in SLE, including statins, has had mixed outcomes to date (Balder et al., 2015). As a result, clear guidelines on monitoring and management of atherosclerosis-associated complications are lacking. Thus, there is an urgent need to improve SLE patient stratification based on CVD profiles to develop adequate therapeutic approaches and reduce the CVD morbidity and mortality in this high-risk population.

In this study, using non-invasive vascular ultrasound imaging to assess the presence of preclinical atherosclerotic plaques, comprehensive global gene expression profiles have been evaluated by RNA sequencing (RNA-seq) on CD14+ circulating monocytes from women with SLE. We identified monocytic transcriptomic profiles associated with the presence of preclinical plaques in women with SLE. Advanced bioinformatic analysis also revealed functional pathways and interactions between the identified differentially expressed genes that could explain the mechanisms leading to accelerated plaque formation. We propose that these molecular signatures could help understand why a subset of women with SLE are predisposed to develop atherosclerosis and are at higher risk of developing clinical CVD.

## Results and Discussion

### Atherosclerosis in the context of SLE is associated with a reprogrammed global monocyte gene expression

We have a well-phenotyped cohort of ~100 atherosclerotic-CVD-free women with SLE scanned for asymptomatic atherosclerotic plaques twice over the course of 5-6 years using non-invasive ultrasound imaging of the carotid and femoral arteries (Smith et al., 2016) (Fig. 1A, Table S1). A subset of patients remained atherosclerotic lesion free over this period (SLE-No Plaque or SLE-NP) whereas some patients persistently showed the presence of plaques during this time (SLE-Plaque or SLE-P). Our work previously demonstrated altered responses in lipid responsive immune-cells, namely invariant natural killer T (iNKT) cells showing increased proliferation and interleukin-4 production associated with the presence of asymptomatic atherosclerosis in SLE (Smith et al., 2016). In addition, we found this altered anti-inflammatory iNKT cell phenotype resulted from the crosstalk between iNKT cells and monocytes (Smith et al., 2016). Monocytes and macrophages play a key role in atherosclerosis plaque formation (Tabas & Lichtman, 2017) and SLE (Ma et al., 2019) pathogenesis. Thus, to further explore the monocyte molecular programs underlying asymptomatic atherosclerosis in SLE, we next investigated the transcriptomic profiles of monocytes in women with SLE with or without atherosclerosis.

**Figure 1.**
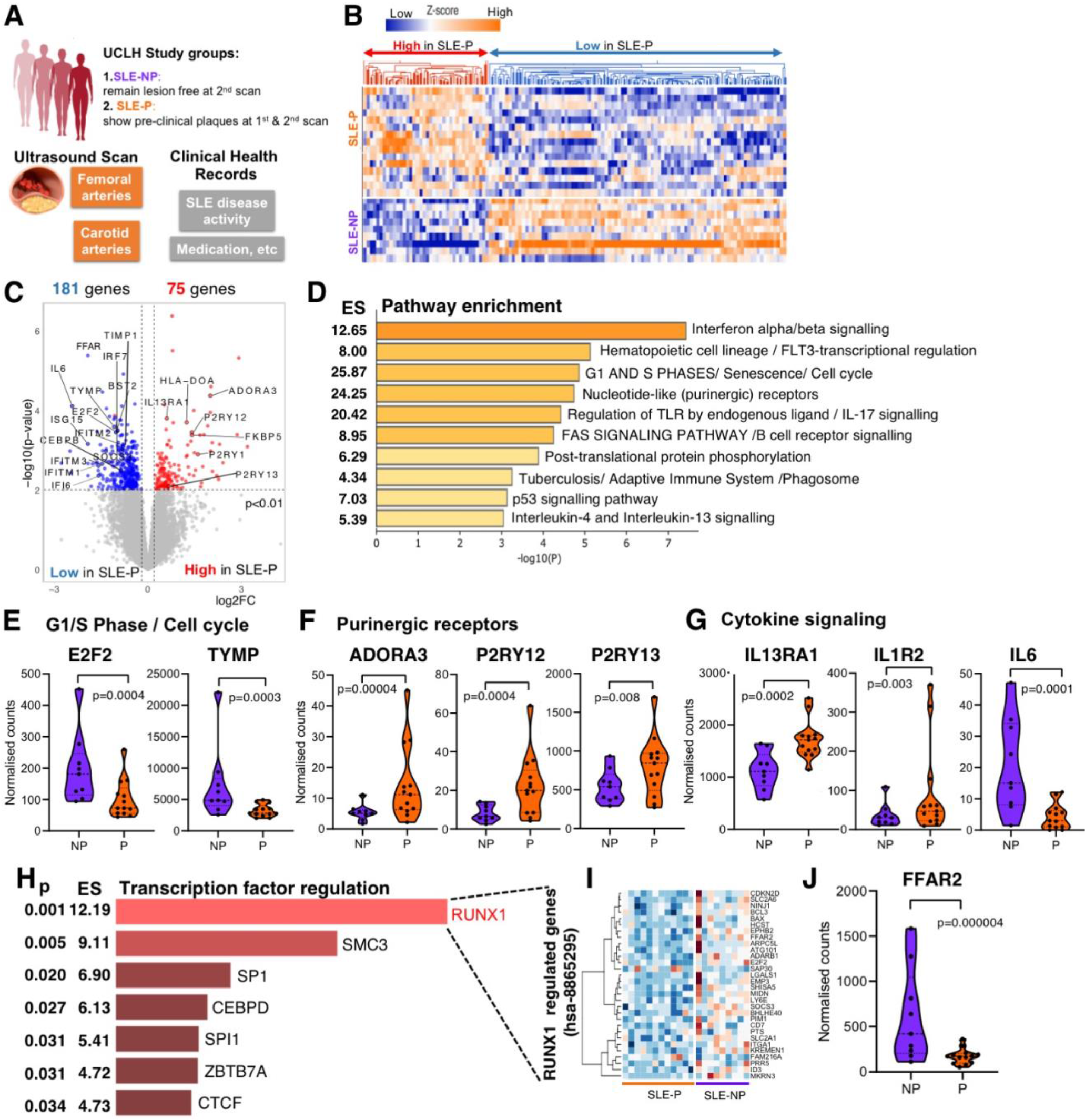
Women with SLE and pre-clinical atherosclerosis (SLE-P) show distinctive patterns of gene expression compared to those without atherosclerosis (SLE-NP) **A**. Summary of study groups. **B**. Clustered heatmap of CD14+ monocyte normalized gene counts showing genes differentially expressed (DE) genes (≥1.5 fold change, p < 0.01) analysed by RNA-seq (SLE-P, n=15; SLE-NP, n=9). **C**. Volcano plot illustrating DE genes clustered in B. **D**. Pathway enrichment analysis of DE genes plotted by significance (-log10 p-value) with enrichment scores (ES) shown. Annotations refer to KEGG, Reactome and Canonical derived pathways. **E-G**. Violin plots for representative genes in the indicated pathways of the normalized gene counts. Statistical analysis from Deseq2 DE analysis shown. Bar represents the median. **H**. Enrichment analysis (EnrichR: ChEA and ENCODE) of transcription factors associated with DE genes in B & D. **I**. Heatmap of normalized gene counts (natural log x+1 transformed) for RUNX1 associated genes (n=30) in SLE-P and SLEP-NP groups. **J**. Violin plot of the normalized gene counts for most significant DE gene in the RUNX1-regulated pathways. Statistical analysis from Deseq2 analysis shown. Bar represents the median.

RNA-seq analysis revealed significant genome wide changes in gene expression (Fig. 1B,C) with a distinctive group of genes significantly induced (75) or reduced (181) in SLE-P compared to those with no plaques (SLE-NP). KEGG/Reactome pathway analysis using Metascape identified interferon signalling (Fig. 1D), cell cycle (Fig.1D, E), purinergic receptor signalling (Fig.1D,F) and cytokine-driven pathways (Fig. 1 D, G) to be differentially enriched (Table 1). These pathways have been implicated in SLE and atherogenesis (Flynn et al., 2019) or both. For instance, local proliferation of macrophages within the plaque has been implicated in the maintenance of macrophage plaque content, particularly in advanced lesions (Hilgendorf et al., 2013). In addition, elevated numbers of circulating monocytes have been associated with increased CVD risk (Coller, 2005) and monocytosis has been causally linked to accelerated atherosclerosis progression and impaired lesion regression (Swirski et al., 2007; Waterhouse et al., 2008). Perhaps counterintuitively, SLE-P monocytes expressed lower levels of genes implicated in cell cycle progression, such as the transcription factor E2F2 (−2.04-fold, P=3.6E-04) (Fig. 1E), typically associated with proliferative malignancies (Cobrinik, 1996; DeGregori, 2002). Interestingly, to our knowledge no reports linking this transcription factor with SLE or atherogenesis in the context of SLE exist. E2F2 was shown to modulate endothelial cell angiogenic responses through G1/S-phase gene expression and proliferation (J. Zhou et al., 2013). We previously reported that gene targets of this family of transcription factors are induced in macrophages from experimental models showing enhanced atherosclerosis (Gage et al., 2018). Further investigations will need to be conducted to assess the role of E2F2 on monocyte biology in the context of atherosclerosis formation in SLE.

**Table 1:**
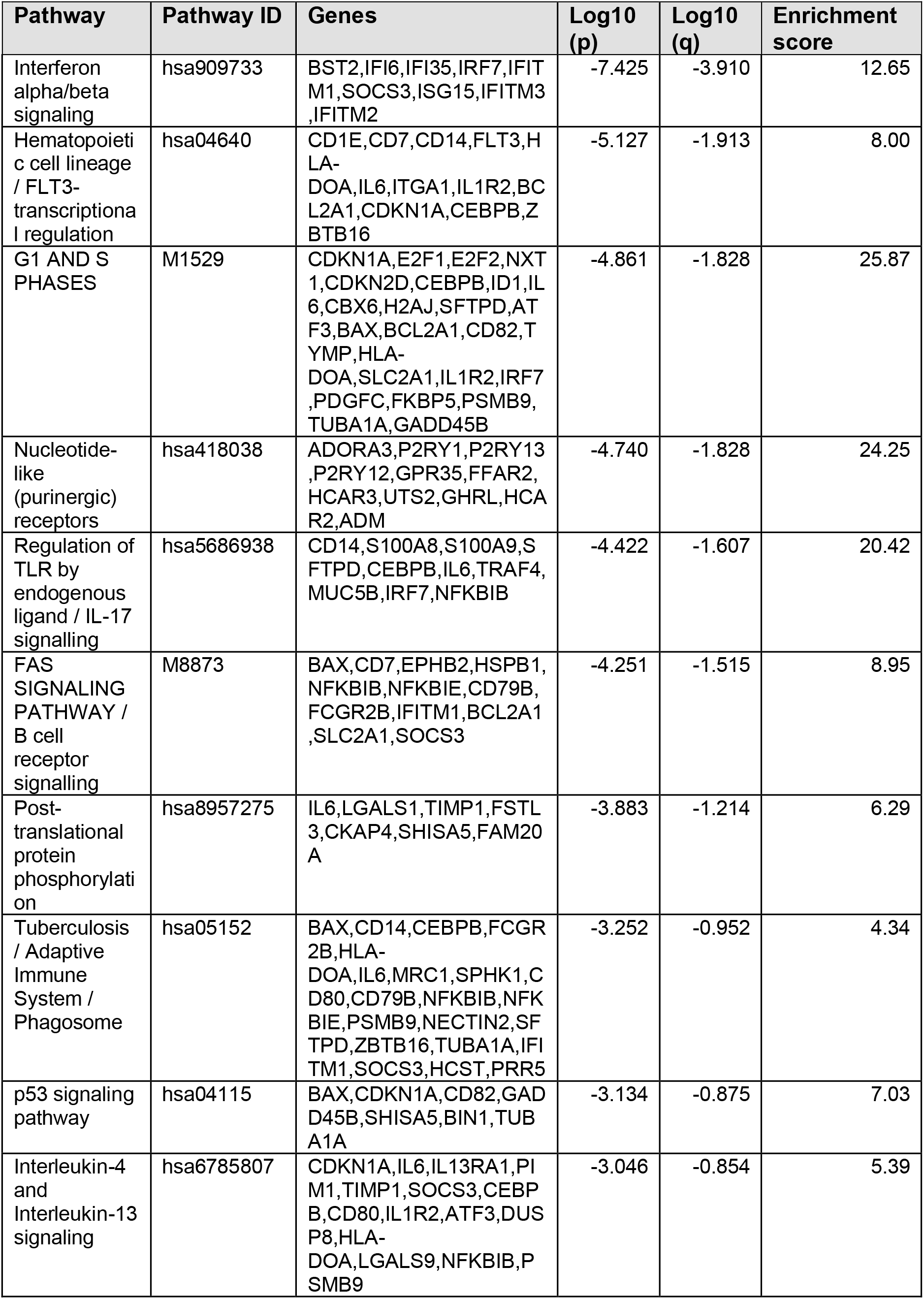
Metascape analysis of enriched Reactome/KEGG/Canonical/Corum pathways in differentially expressed genes in SLE-P vs SLE-NP monocytes. Top 10 clusters with representative enriched terms (Pathway) and KEGG Functional Sets, KEGG Pathway, Reactome Gene Sets, Canonical Pathways, CORUM, TRRUST, DisGeNET and PaGenBase annotation identifier (Pathway ID). Genes include all genes enriched in pathway from differential gene expression seed list. Log_10_(p) refers to the hypergeometric distribution test statistic in log base 10. Log_10_(q) refers to the multi-test adjusted p-value in log base 10. Enrichment score indicates how many fold more pathway members belong to a given pathway than expected by chance.

Purinergic signalling complexes, including extracellular nucleotides and their receptors such as the P2 receptors (P2RY12), induced in the SLE-P monocytes (Fig.1D,F), are considered major regulatory systems that can regulate the development and course of multiple immune-mediated inflammatory diseases such as SLE (Antonioli et al., 2019). Previously, the P2X7 receptor, as part of the NLRP3 inflammasome signalling, has been implicated in the pathogenesis of SLE and other autoimmune diseases (Cao et al., 2019) as well as in atherosclerosis formation (Wallentin et al., 2009). Beyond P2X7R, the P2RY12 agonist Ticagrelor, known to attenuate platelet-leukocyte interactions (Schrottmaier et al., 2015), has been reported to reduce cardiovascular events in CVD patients, dampen vascular dysfunction and reduce atherosclerosis in experimental models of disease by inhibiting endothelial cell inflammation (Ganbaatar et al., 2018). More recently, P2RY12, better known as a microglia marker (Haage et al., 2019), was implicated in promoting vascular smooth muscle cell foam cell formation through the inhibition of autophagy (Pi et al., 2020), however its impact on monocyte function is not well defined. Additionally, an agonist for P2RY13 increases the uptake of HDL-cholesterol (HDL-C) by the liver in experimental models of atherosclerosis (Goffinet et al., 2014) and has been shown to protect against atherosclerosis through its role on the anti-atherogenic reverse cholesterol transport (Lichtenstein et al., 2015). Whether these actions are important for monocyte function in the context of preclinical atherosclerosis is unknown.

In addition to these, other pathways involved in cytokine and toll-like receptor signalling, known to be implicated in atherogenesis and autoimmune diseases (Hoeksema et al., 2012; Li et al., 2020; Ryu et al., 2019) are enriched in SLE-P monocytes (Fig.1D,G). For instance, IL13RA1, one of the subunits of the receptor for the immunomodulatory cytokine IL-13, observed at higher levels in SLE patients (Morimoto et al., 2001), as well as IL-6, which contributes to chronic inflammation and autoimmunity (Tanaka et al., 2014). Additionally, the anti-IL-6 receptor antibody Tocilizumab has proven effective in patients with rheumatoid arthritis (Nishimoto et al., 2004). Whereas, IL-1R1 signalling has been shown to induce human Th17 cell differentiation (Fleischmann et al., 2003; Sha & Markovic-Plese, 2011). The identified gene expression profiles of monocytes associated with the presence of preclinical atherosclerosis suggest a less inflammatory status (Fig.1D,G), in support of our previous findings in iNKT cells (Smith et al., 2016). Interestingly, recent reports have revealed the existence of a subset of “foamy” macrophages engorged with lipids showing an anti-inflammatory profile within atherosclerotic plaques (see below) (Willemsen & Winther, 2020).

To further investigate mechanisms explaining atherosclerosis plaque formation in SLE, we used the EnrichR algorithm to identify transcription factors involved in the regulation of the differentially (DE) genes in Fig. 1B,C. This analysis highlighted a potential role for RUNX1, a transcription factor implicated in hematologic malignancies (Olofsen & Touw, 2020) but not previously related to atherosclerosis (Fig. 1H, Table 2). Interestingly, a number of known RUNX1-regulated targets show reduced expression in SLE-P monocytes (Fig. 1I). One of the most differentially expressed genes between the SLE-P and SLE-NP groups in this subset is the free fatty acid receptor 2 or FFAR2/GPR43 (Kimura et al., 2020), a G-protein coupled receptor activated by short-chain fatty acids (SCFAs), mainly acetate, butyrate and propionate (Fig. 1J). These SCFAs have shown immunomodulatory roles in monocytes by reducing the production of tumour necrosis factor alpha and monocyte chemotactic protein-1 while increasing the production of prostaglandin E2 (Cox et al., 2009). In addition, FFAR2 signalling has been demonstrated to inhibit nuclear translocation of NF-kB, thereby reducing the expression of inflammatory cytokines (IL-1 and IL-6) in cultured cells (Lee et al., 2013) (Ang et al., 2016).

**Table 2:**
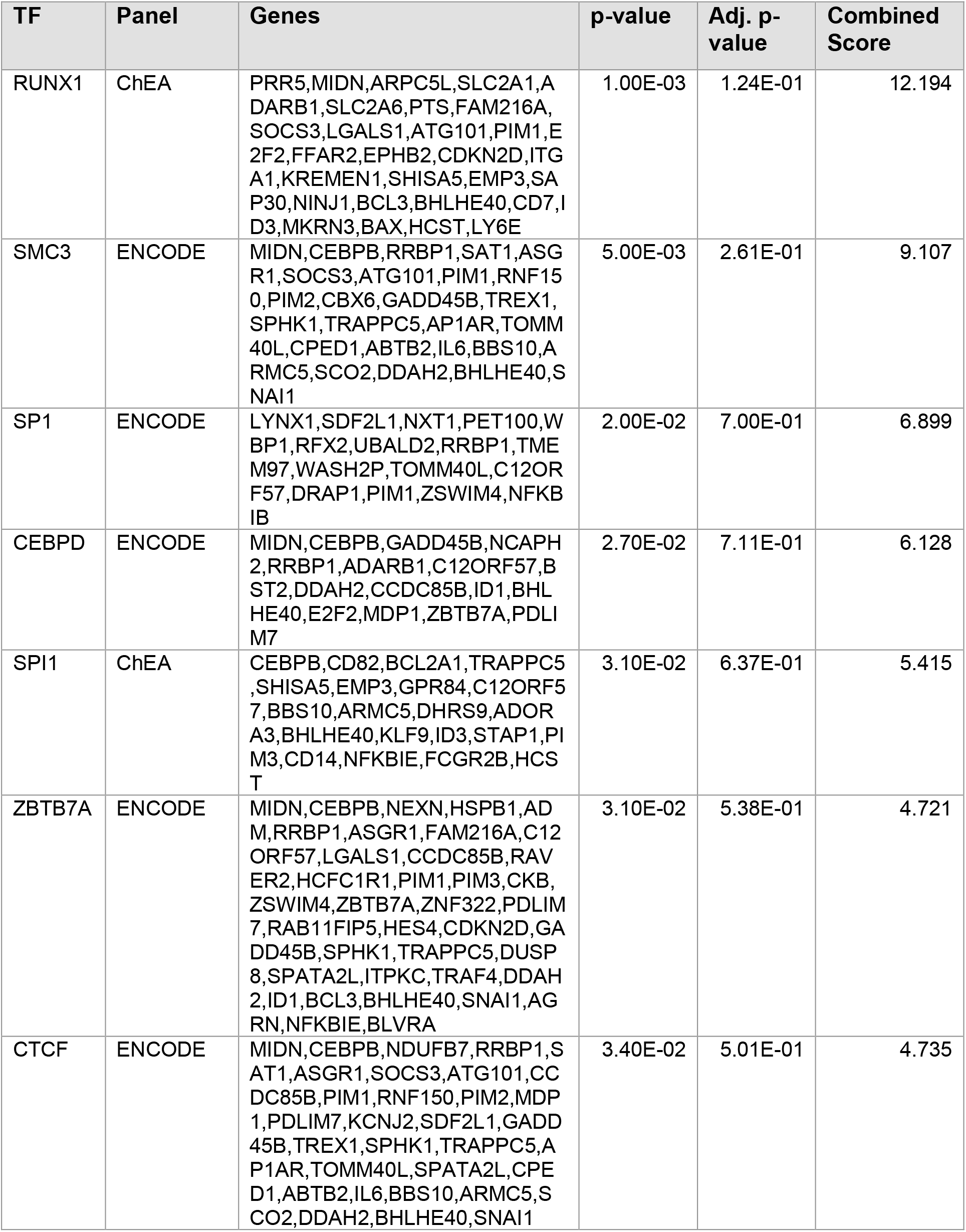
EnrichR analysis of transcription factors (TFs) regulating differentially expressed genes in SLE-P vs SLE-NP monocytes. Table of transcription factors and consensus target genes using ENCODE and ChEA. TF refers to the transcription factor identifier. Panel refers to either ENCODE or ChEA repositories used to enrich term. Genes includes all genes from input seed gene list targeted by specific transcription factor. P-value is Fisher’s exact test statistic. Adj. p-value refers to the adjusted p-value using Benjamini-Hochberg correction for multiple hypothesis testing. Combined score refers to the p-value and z-score (deviation from expected rank) calculated by multiplying the two values (combined score = natural log(p-value) multiplied by z-score).

Overall, we have identified novel pathways and genes associated with the presence of atherosclerotic plaques in women and confirmed the implication of pathways already associated with the development of atherosclerotic plaques.

### Interferon signatures do not distinguish the presence of atherosclerosis in women with SLE

Attempts to stratify SLE patients according to unique molecular signatures have focused largely on the involvement of the interferon family of cytokines in the pathophysiology of the disease. Specifically, type 1 interferon (IFN-α and IFN-β) glycoproteins, with highly potent antiviral, anti-proliferative and immunoregulatory actions (Rönnblom & Alm, 2001). Activation of IFN-α, which shows elevated circulating levels in SLE patients (Hooks et al., 1979; Ytterberg & Schnitzer, 1982), has long been considered to play a key role in the disease. In addition, high levels of IFN-α have been shown to associate with SLE disease activity (Baechler et al., 2003; Bengtsson et al., 2000; Crow, 2014; Kanayama et al., 1989; Obermoser & Pascual, 2010). The term “IFN-signature” has traditionally been used to describe the overexpression of IFN-I-induced transcripts and evidence of IFN-I signalling (Bennett et al., 2003) in SLE patients compared to healthy controls. This is mainly because serum concentrations of IFN-α are difficult to be reliably measured. Instead, gene expression levels of IFN-α regulated genes are frequently used to monitor IFN activity in SLE to deduce an “IFN score” (El-Sherbiny et al., 2018; Figgett et al., 2019; Yao et al., 2009). However, existing reports have evidenced only about 50% SLE patients show an activated IFN signature (Baechler et al., 2003).

Type I IFN signalling was shown to be dysregulated in symptomatic atherosclerotic plaque macrophages (Chai et al., 2018; Fernandez et al., 2019), however, two of the reported SLE interferon signatures (El-Sherbiny et al., 2018; Figgett et al., 2019) do not appear to stratify the presence of atherosclerosis in the women in our cohort (Fig.2A,B). Pathway analysis did indeed identify type I IFN signalling as one of the most enriched pathways in monocytes from the SLE-P group (Fig. 1D; Table 3). Nevertheless, the IFN-regulated differentially expressed genes identified show minimal overlap with those previously reported in (El-Sherbiny et al., 2018; Figgett et al., 2019) (not shown) and, could also not distinguish the presence of atherosclerosis (Fig.2C). Strikingly, the interferon signature in SLE-P monocytes was significantly down-regulated compared to SLE-NP monocytes (Fig. 2D)

**Figure 2.**
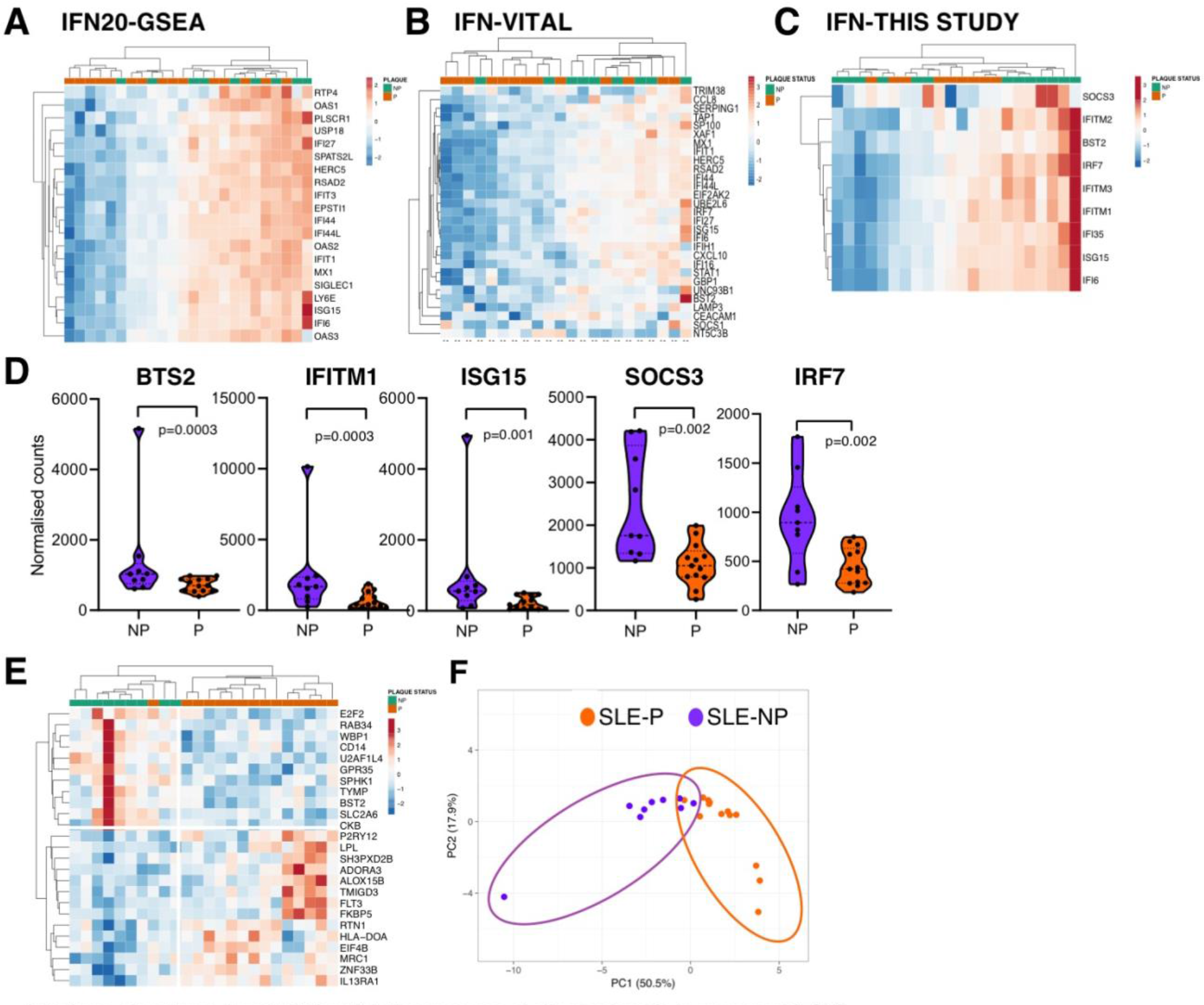
Interferon signatures do not distinguish the presence of atherosclerosis in women with SLE. **A-C**. Unsupervised clustering heatmaps of CD14+ monocyte normalized counts in SLE-P vs SLE-NP groups of the indicated IFN signatures: A, IFN20-GSEA (Ay et al., 2020); B, IFN-Vital (Vital et al., 2018); C, IFN signature identified in Fig. B,E. **D**. Violin plots for representative genes in the IFN signature shown in C showing the normalized gene counts in SLE-P (orange) and SLE-NP (purple). Statistical analysis from Deseq2 DE analysis shown. Bar represents the median. **E**. Unsupervised clustering heatmap of top significant DE genes between SLE-P vs SLE-NP groups showing better clustering of the presence of atherosclerosis. **F**. Principal component analysis (PCA) of top significant DE genes shown in E. PCA X and Y axis show principal component 1 and 2 that explain 50.5% and 17.9% of the total variance, respectively.

**Table 3:** Differentially expressed genes in SLE-P monocytes in the IFNab signalling pathway. Table of differential gene expression values for genes enriched in IFNab signalling pathway (hsa909733) in SLE-P compared to SLE-NP. Fold change refers to the direction of change in gene expression with negative numbers depicting downregulated gene expression in linear format.

| **Genes** | **Annotation** | **p-value** | **Fold change** |
| --- | --- | --- | --- |
| BST2 | bone marrow stromal cell antigen 2 | 3.00E-04 | -1.96 |
| ISG15 | ISG15 ubiquitin like modifier | 1.00E-03 | -1.75 |
| IFITM2 | interferon induced transmembrane protein 2 | 1.00E-03 | -3.79 |
| IRF7 | interferon regulatory factor 7 | 2.00E-03 | -1.96 |
| SOCS3 | suppressor of cytokine signaling 3 | 2.00E-03 | -2.35 |
| IFITM3 | interferon induced transmembrane protein 3 | 2.00E-03 | -2.77 |
| IFITM1 | interferon induced transmembrane protein 1 | 3.00E-03 | -3.00 |
| IFI6 | interferon alpha inducible protein 6 | 5.00E-03 | -1.76 |
| IFI35 | interferon induced protein 35 | 5.00E-03 | -2.99 |

A better stratification between SLE-P compared to SLE-NP was identified when we included the most significantly differentially expressed genes as shown by unsupervised hierarchical clustering (Fig.2E), and principal component analysis (Fig.2F). These findings may be relevant considering existing clinical trials of therapies targeting type I IFN signalling have shown variable efficacy in the treatment of SLE and other autoimmune diseases (Jiang et al., 2020). Elucidating a more definitive molecular signature of atherosclerosis in the context of SLE will be critical for patient stratification and identifying therapies for the treatment of atherosclerosis in SLE.

### Overlapping and distinct signalling pathways exist between SLE-P monocytes and atherosclerotic plaque macrophages

Recent advances in the application of immunohistochemistry and bulk RNA-seq analysis (Chai et al., 2018) as well as the rapid progress of single-cell sequencing technologies (Fernandez et al., 2019) have now enabled more comprehensive transcriptional mapping of the cell types and their phenotypes present in atherosclerotic plaques. Specific subsets of cells can now be distinguished based on overlapping or differing transcriptomes giving novel insights regarding immune cell function and plaque composition. Such studies have highlighted the presence of subsets of pro-inflammatory, foamy anti-inflammatory or resident-like macrophages (Willemsen & Winther, 2020) in both murine and human plaques. In addition, some of these reports have shown that foamy macrophages are more anti-inflammatory compared to non-foamy macrophages in human atherosclerotic plaques, as foamy macrophages express significantly less Il1b and other pro-inflammatory markers (Kim et al., 2018).

We next interrogated which pathways overlap or are distinctive between the gene expression profiles of circulating monocytes in SLE-P patients in our study, and those of the symptomatic plaque macrophages reported in two independent studies (Fig.3). Only a subset of genes enriched in SLE-P monocytes overlap with genes enriched in plaque macrophages (27 gene overlap with genes from the single cell analysis and 11 gene overlap with those identified in the immunohistochemistry-based study) (Fig.3A, Table 4&5). One of the overlapping genes S100A9, is part of the S1000A8/A9 damage-associated molecular pattern protein heterodimer shown to correlate with the severity of coronary artery disease (Cotoi et al., 2014; Kraakman et al., 2017; Peng et al., 2011). This complex is also emerging as a novel exciting therapeutic target to treat monocytosis and atherosclerosis in metabolic diseases (Kraakman et al., 2017; Xiao et al., 2020), as it is thought the increase in circulating monocytes could contribute to plaque macrophage accumulation and atherosclerosis (Cotoi et al., 2014). Of note, both genes in the complex show reduced expression in the SLE-P monocytes compared to those in SLE-NP (S100A9: −1.68-fold, P=3.8E-03; S100A8: −1.77-fold, P=2.4E-03) perhaps reflecting the asymptomatic or preclinical stage of the disease in these SLE patients or differences in metabolic/inflammatory context.

Further Metascape analysis revealed that the limited number of common genes had a large degree of functional overlap with the carotid plaque macrophage transcriptomes as demonstrated by the light blue overlapping lines on the circos plot (Fig.3B). Subsequent analysis of these overlapping pathways (Table 6) confirmed the relevance of signalling by interleukins, including anti-inflammatory cytokines like IL-13 and IL-10, further supporting our previous reports (Smith et al., 2016) (Fig.3C). Interestingly, IL-13 has been shown to reduce the expression of CD36 receptor responsible for oxidised lipid uptake in macrophages and subsequent foam cell formation (Yakubenko et al., 2013). In addition, targeting IL-13 signal transduction pathways has been proposed to ameliorate vascular dysfunction in experimental models of lupus (Furumoto et al., 2017) and a number of therapeutics targeting IL-13 are being assessed in clinical trials against various autoimmune diseases (Mao et al., 2019). Furthermore, Toll-like receptor signalling and adaptive immune responses are enriched (Fig.3C). Collectively, these findings show a substantial overlap in the pathways relevant in circulating monocytes in SLE patients with preclinical atherosclerosis and those in symptomatic atherosclerosis plaques.

**Figure 3.**
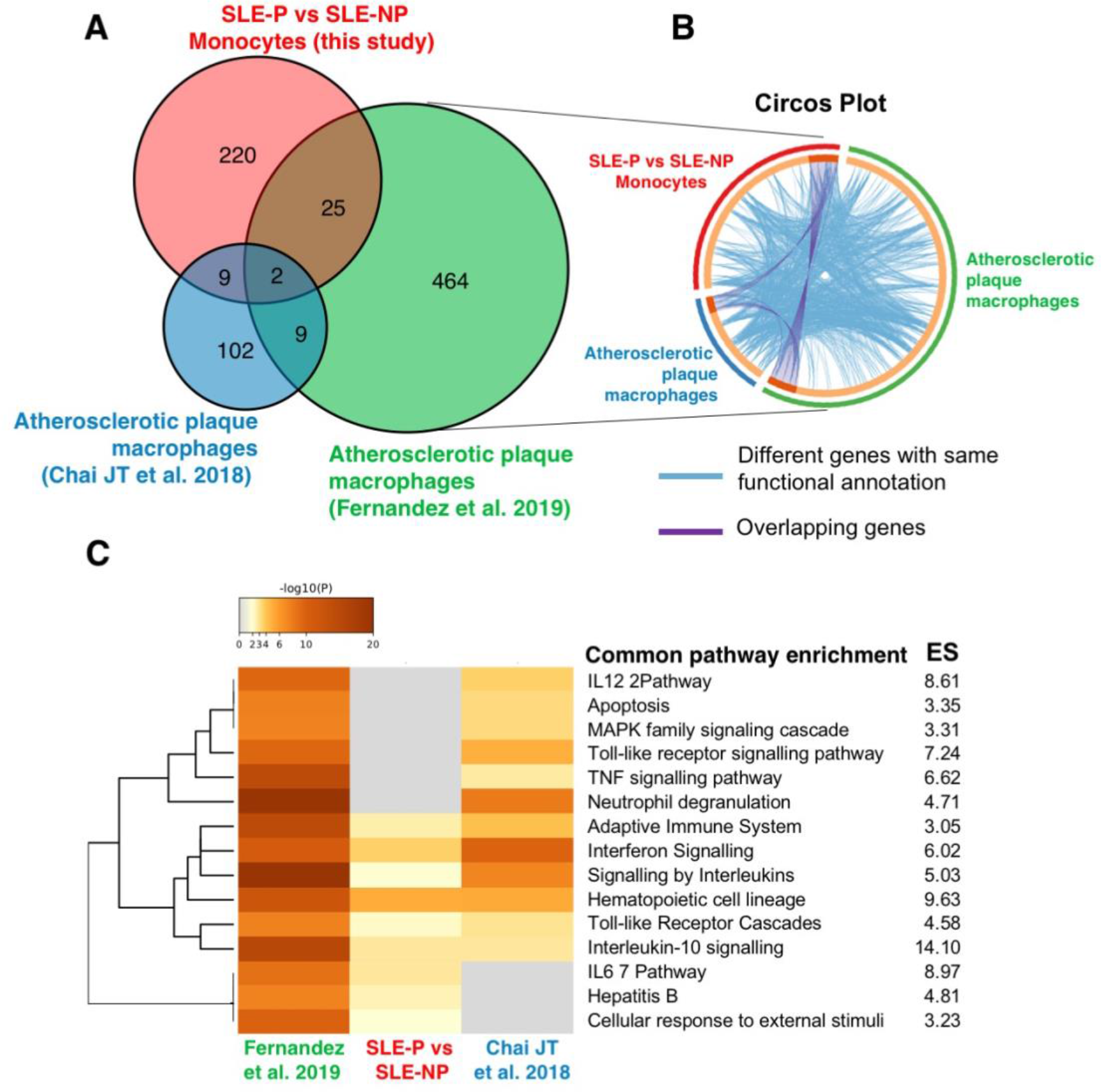
Overlapping signalling pathways exist between SLE-P monocytes and reported atherosclerotic plaque macrophages. **A**. Venn diagram comparing the DE genes identified Fig. **1B** (red), enriched genes in asymptomatic plaque macrophages (Fernandez et al. 2016) (green) and (Chai JT et al. 2018) (blue). Numbers represent the number of genes in each overlapping or non-overlapping group. **B**. Circos plot highlighting the overlap between the indicated datasets: Blue lines link DE genes sharing the same gene ontology, whereas purple lines link overlapping genes. **C**. Pathway enrichment analysis of common DE genes plotted by significance (-Iog10 p-value) with enrichment scores (ES) shown. Annotations refer to KEGG, Reactome and Canonical derived pathways.

**Table 4.**
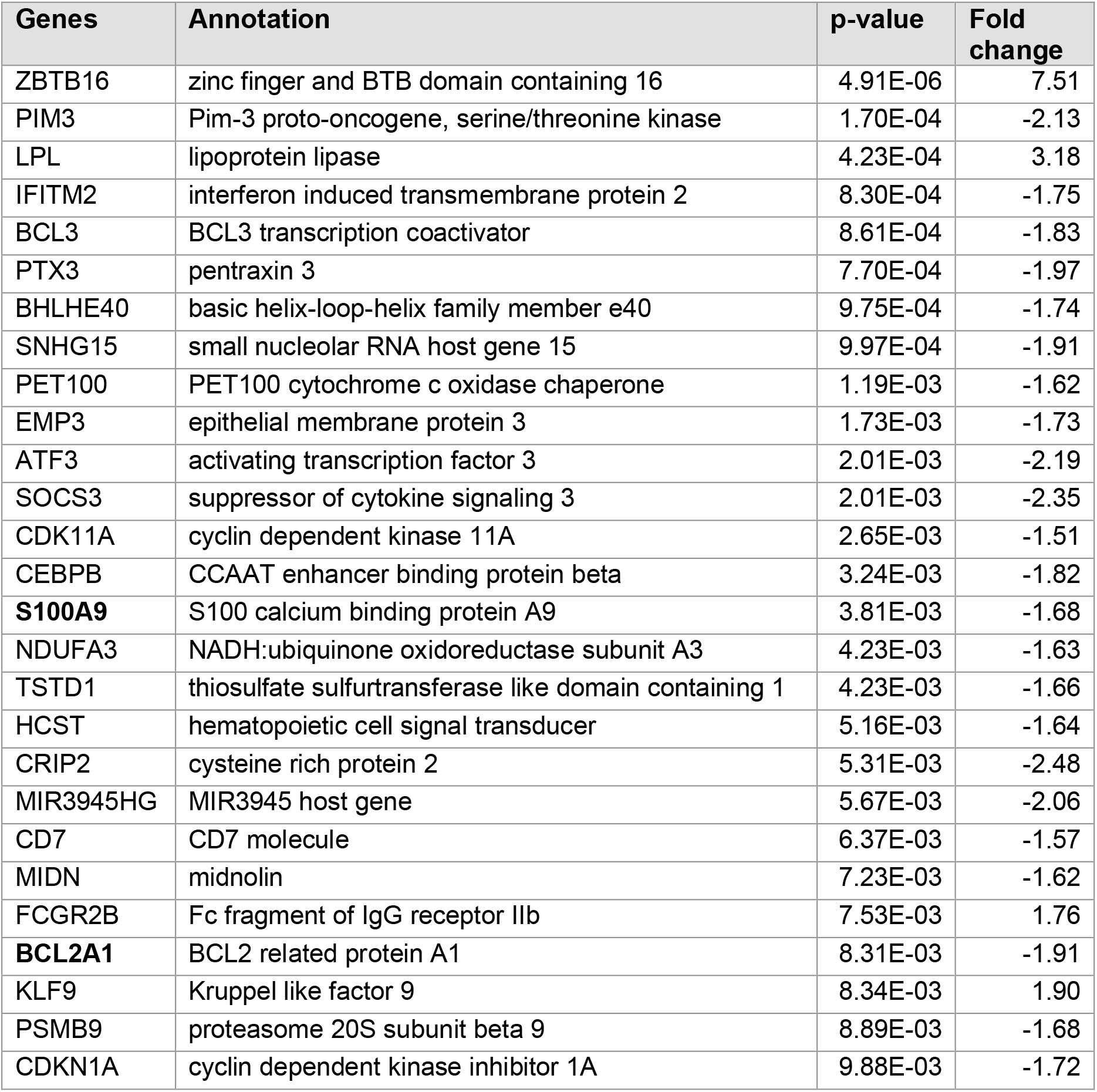
Genes differentially expressed in SLE-P monocytes and symptomatic plaque macrophages identified by scRNA-seq analysis (Fernandez et al., 2019) Fold change as in Table 3. Genes in bold also conserved in laser capture microdissection symptomatic plaque macrophage expression list.

**Table 5.** Genes differentially expressed in SLE-P monocytes and laser capture microdissection symptomatic plaque macrophages (Chai et al., 2018) Fold change as in Table 3. Genes in bold also conserved in scRNA-seq symptomatic plaque macrophage expression list.

| **Genes** | **Annotation** | **p-value** | **Fold change** |
| --- | --- | --- | --- |
| ISG15 | ISG15 ubiquitin like modifier | 7.09E-04 | -3.79 |
| SAP30 | Sin3A associated protein 30 | 1.17E-03 | 2.80 |
| P2RY1 | purinergic receptor P2Y1 | 1.29E-03 | 3.03 |
| ASGR1 | asialoglycoprotein receptor 1 | 1.90E-03 | -1.59 |
| S100A8 | S100 calcium binding protein A8 | 2.37E-03 | -1.77 |
| IL4I1 | interleukin 4 induced 1 | 3.45E-03 | -1.85 |
| **S100A9** | S100 calcium binding protein A9 | 3.81E-03 | -1.68 |
| IFI6 | interferon alpha inducible protein 6 | 4.51E-03 | -2.99 |
| ADM | adrenomedullin | 4.83E-03 | -1.61 |
| **BCL2A1** | BCL2 related protein A1 | 8.31E-03 | -1.91 |
| KCNJ2 | potassium inwardly rectifying channel subfamily J member 2 | 9.35E-03 | 2.04 |

**Table 6.**
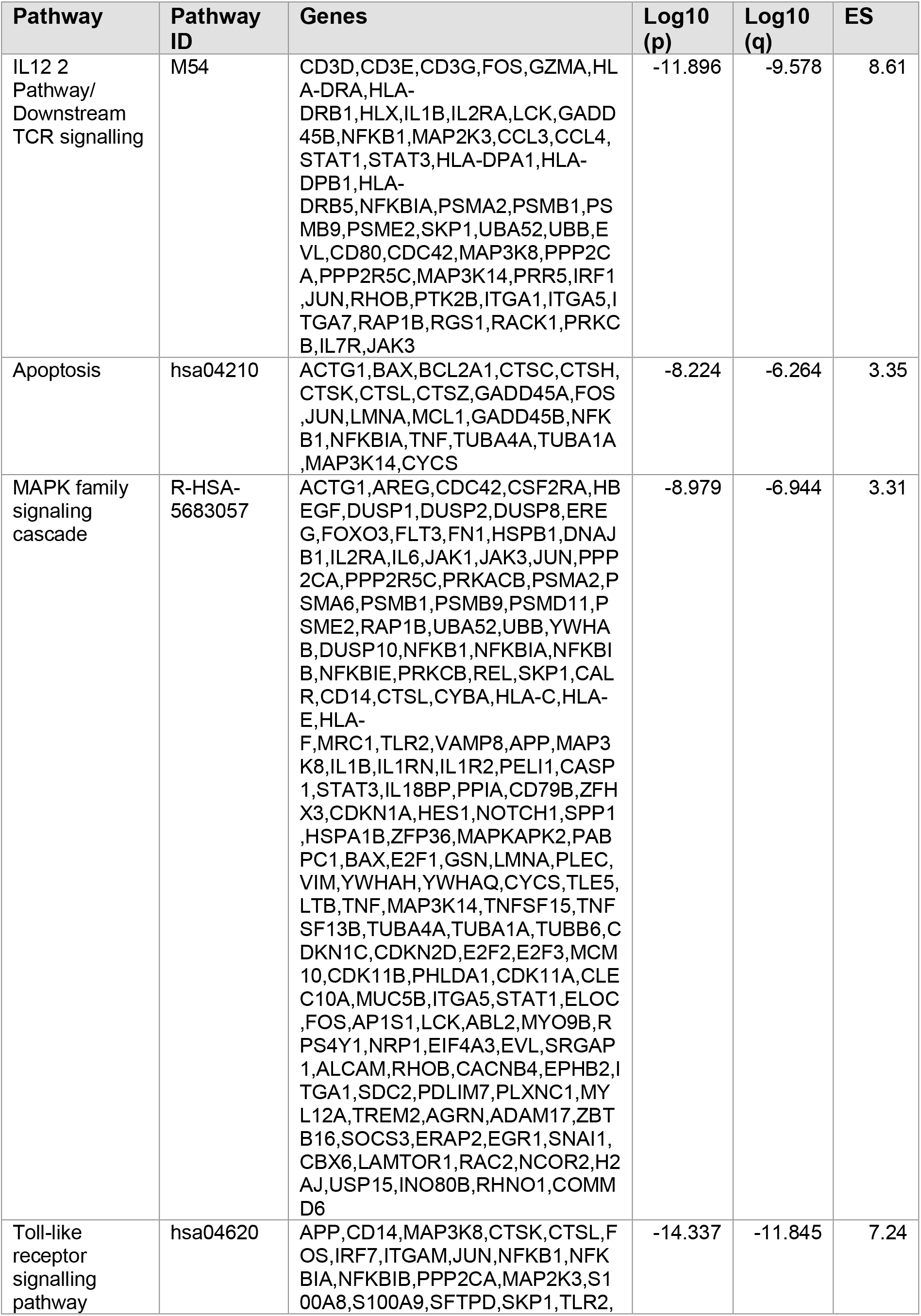
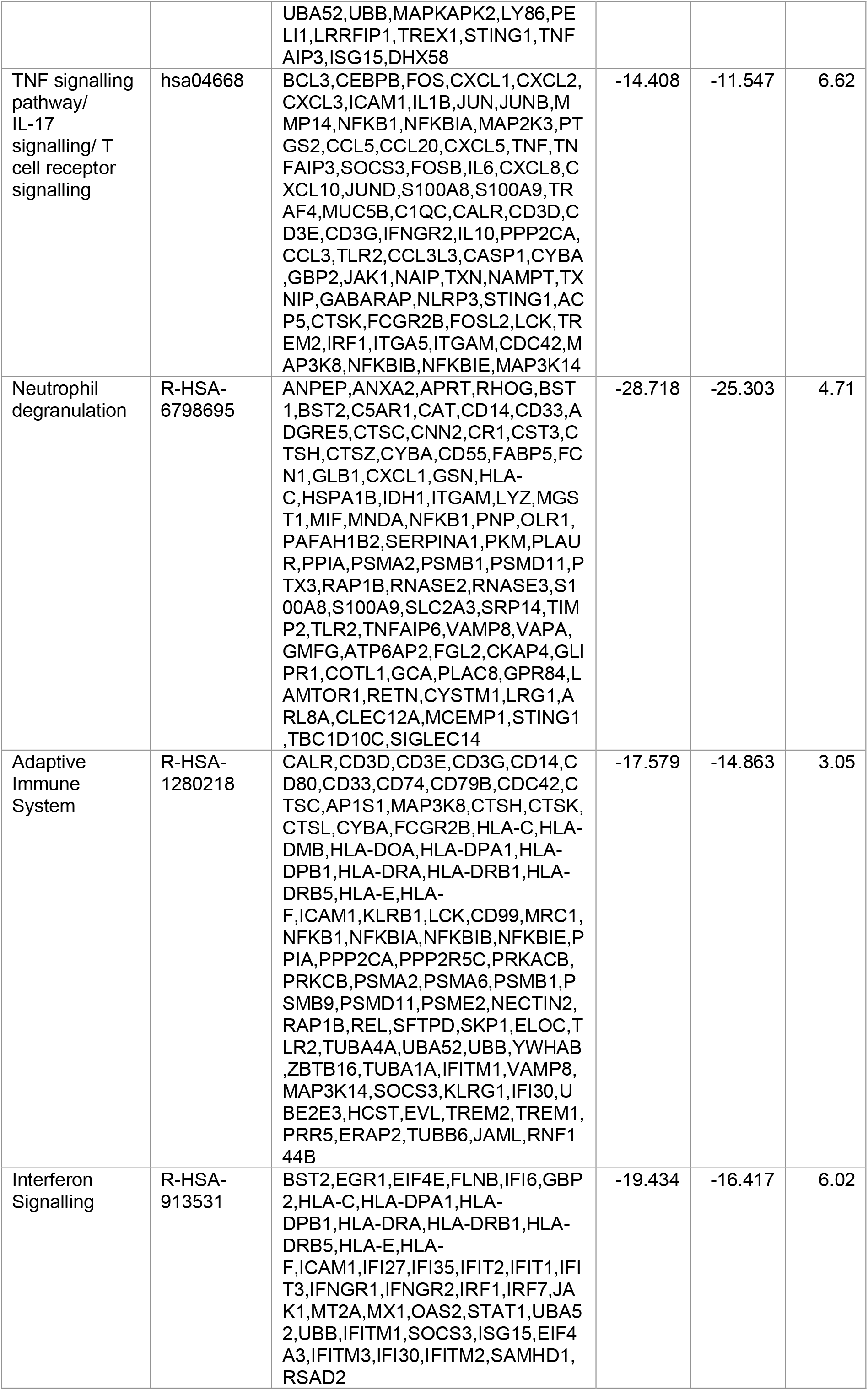
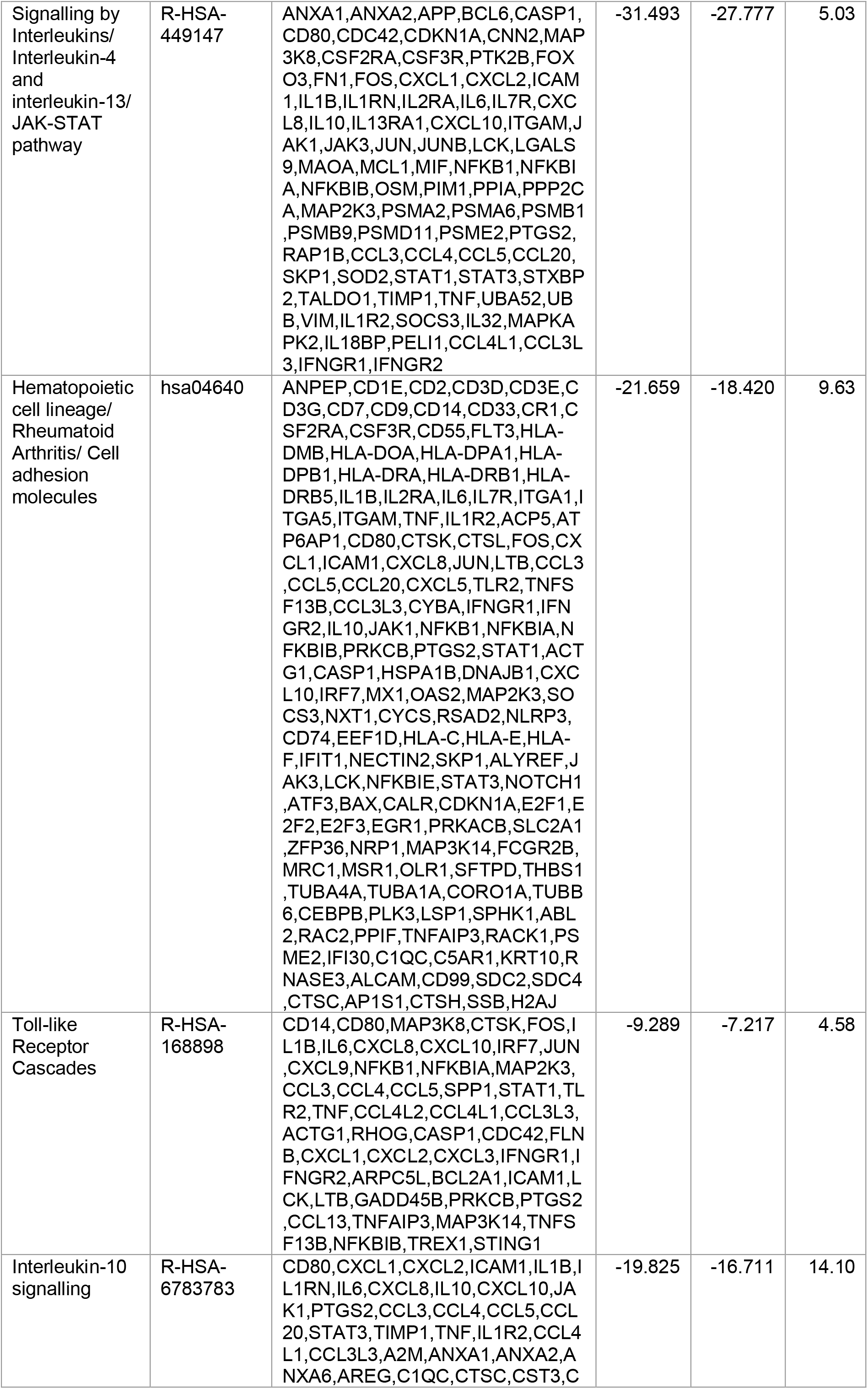
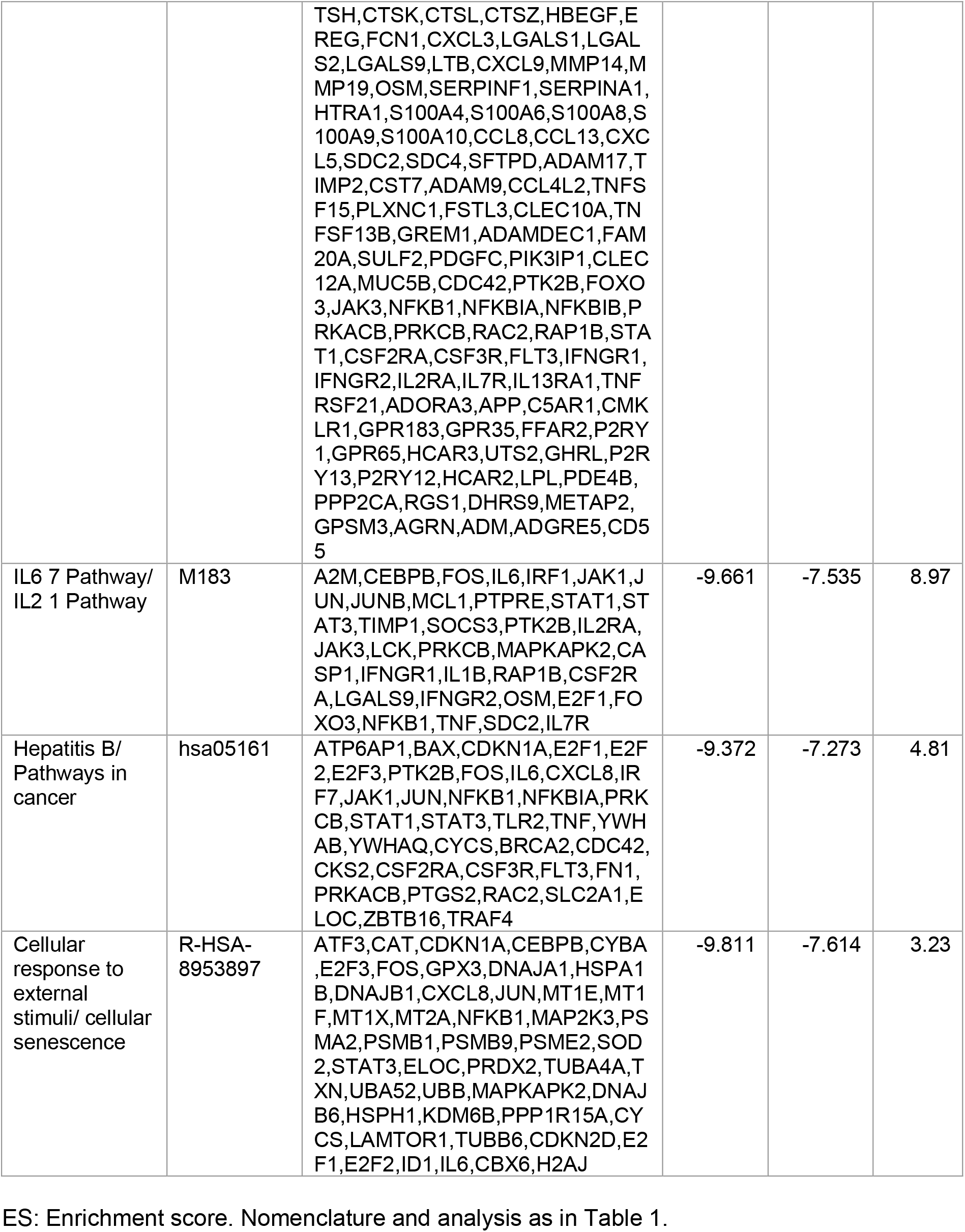
Overlapping pathways and enrichment across SLE-P monocytes, scRNA-seq (Fernandez et al., 2019) and laser capture microdissection (Chai et al., 2018) symptomatic plaque macrophages.

### Enhanced interaction network analysis identifies pathways involved in the adaptive immune system

We next aimed to understand the pathways associated with atherosclerosis in SLE based on known protein-protein interactions (PPI). As a seed list, we selected genes differentially expressed between SLE-P and SLE-NP at a stringent significance threshold of p<0.001 (Table 7) and visualised how these mapped onto the STRINGdb PPI-network using Network Analyst (Fig. 4A) and Reactome (Fig. 4D). Within the protein network, smaller clusters of densely connected regions, known as modules, reflect functional associations between biological processes of the proteins characterised. The initial PPI-network interacting with the seed list predicted 3ii nodes (proteins) and 331 edges (interactions) (24 genes were excluded due to not being present in STRINGdb PPI-network reference database at the inclusion threshold). This analysis identified a number of relevant nodes including ISG15, a well-known IFN-regulated gene, FLT3 (an oncogenic receptor tyrosine kinase implicated in hematological disorders) (Voisset et al., 2020) and EIF4B, the translation eukaryotic initiation factor involved in the mTOR pathway and translation of interferon-regulated genes (Joshi et al., 2010).

**Table 7:** Differentially expressed genes in SLE-P monocytes DIAMOnD seed set.

| Seeds | Annotation | p-value | Fold change |
| --- | --- | --- | --- |
| ZNF33B | zinc finger protein 33B | 4.00E-07 | 1.71 |
| ZBTB16 | zinc finger and BTB domain containing 16 | 5.00E-06 | 7.51 |
| ZNF302 | zinc finger protein 302 | 3.00E-06 | 1.72 |
| FFAR2 | free fatty acid receptor 2 | 4.00E-06 | -3.79 |
| U2AF1L4* | U2 small nuclear RNA auxiliary factor 1 like 4 | 1.00E-05 | -1.73 |
| TMIGD3 | transmembrane and immunoglobulin domain containing 3 | 3.00E-05 | 4.05 |
| WBP1 | WW domain binding protein 1 | 4.00E-05 | -2.75 |
| ADORA3 | adenosine A3 receptor | 4.00E-05 | 3.99 |
| ADARB1 | adenosine deaminase RNA specific B1 | 8.00E-05 | -1.79 |
| IL6 | interleukin 6 | 8.00E-05 | -5.38 |
| SCO2 | synthesis of cytochrome C oxidase 2 | 9.00E-05 | -2.51 |
| MRC1 | mannose receptor C-type 1 | 1.00E-04 | 2.77 |
| FLT3 | fms related receptor tyrosine kinase 3 | 1.00E-04 | 4.00 |
| RNF150 | ring finger protein 150 | 1.00E-04 | 61.26 |
| IL13RA1 | interleukin 13 receptor subunit alpha 1 | 2.00E-04 | 1.52 |
| DRAP1 | DR1 associated protein 1 | 2.00E-04 | -1.82 |
| DDAH2 | dimethylarginine dimethylaminohydrolase 2 | 2.00E-04 | -1.87 |
| ABTB2 | ankyrin repeat and BTB domain containing 2 | 2.00E-04 | -2.09 |
| PIM3 | Pim-3 proto-oncogene, serine/threonine kinase | 2.00E-04 | -2.13 |
| SPHK1 | sphingosine kinase 1 | 1.00E-04 | -2.34 |
| HLA-DOA | major histocompatibility complex, class II, DO alpha | 2.00E-04 | 2.36 |
| GAPT | GRB2 binding adaptor protein, transmembrane | 2.00E-04 | 1.75 |
| CD14 | CD14 molecule | 2.00E-04 | -1.57 |
| HLX | H2.0 like homeobox | 2.00E-04 | -1.79 |
| CCDC122 | coiled-coil domain containing 122 | 2.00E-04 | 3.77 |
| TYMP | thymidine phosphorylase | 3.00E-04 | -2.12 |
| ALOX15B | arachidonate 15-lipoxygenase type B | 4.00E-04 | 3.47 |
| LPL | lipoprotein lipase | 4.00E-04 | 3.18 |
| FKBP5 | FKBP prolyl isomerase 5 | 4.00E-04 | 2.66 |
| P2RY12 | purinergic receptor P2Y12 | 4.00E-04 | 2.65 |
| RRBP1 | ribosome binding protein 1 | 4.00E-04 | -1.60 |
| SLC2A6 | solute carrier family 2 member 6 | 4.00E-04 | -1.77 |
| GPR35 | G protein-coupled receptor 35 | 4.00E-04 | -1.79 |
| NFKBIE | NFKB inhibitor epsilon | 4.00E-04 | -1.86 |
| BST2 | bone marrow stromal cell antigen 2 | 3.00E-04 | -1.96 |
| PLEKHN1 | pleckstrin homology domain containing N1 | 3.00E-04 | -1.97 |
| E2F2 | E2F transcription factor 2 | 4.00E-04 | -2.04 |
| ITGA7 | integrin subunit alpha 7 | 3.00E-04 | -2.46 |
| CKB | creatine kinase B | 3.00E-04 | -3.09 |
| TPST1 | tyrosylprotein sulfotransferase 1 | 7.00E-04 | 4.90 |
| CTTNBP2 | cortactin binding protein 2 | 7.00E-04 | 4.16 |
| CD1E | CD1e molecule | 6.00E-04 | 2.60 |
| SH3PXD2 B | SH3 and PX domains 2B | 8.00E-04 | 2.42 |
| ZNF583 | zinc finger protein 583 | 9.00E-04 | 1.66 |
| RTN1 | reticulon 1 | 6.00E-04 | 1.60 |
| AP1AR | adaptor related protein complex 1 associated regulatory protein | 7.00E-04 | 1.54 |
| EIF4B | eukaryotic translation initiation factor 4B | 7.00E-04 | 1.53 |
| RAB34* | RAB34, member RAS oncogene family | 9.00E-04 | -1.52 |
| CIB1 | calcium and integrin binding 1 | 8.00E-04 | -1.56 |
| NCAPH2 | non-SMC condensin II complex subunit H2 | 6.00E-04 | -1.64 |
| BLVRA | biliverdin reductase A | 8.00E-04 | -1.72 |
| CD82 | CD82 molecule | 7.00E-04 | -1.73 |
| IFITM2 | interferon induced transmembrane protein 2 | 8.00E-04 | -1.75 |
| ANKRD9 | ankyrin repeat domain 9 | 8.00E-04 | -1.76 |
| BCL3 | BCL3 transcription coactivator | 9.00E-04 | -1.83 |
| LGALS1 | galectin 1 | 7.00E-04 | -1.93 |
| BIN1 | bridging integrator 1 | 9.00E-04 | -1.94 |
| PTX3* | pentraxin 3 | 8.00E-04 | -1.97 |
| RNASE3 | ribonuclease A family member 3 | 8.00E-04 | -2.23 |
| ISG15 | ISG15 ubiquitin like modifier | 7.00E-04 | -3.79 |
| ATG101 | autophagy related 101 | 1.00E-03 | -1.58 |
| BHLHE40 | basic helix-loop-helix family member e40 | 1.00E-03 | -1.74 |
| SLC2A1 | solute carrier family 2 member 1 | 1.00E-03 | -1.78 |
| NECTIN2 | nectin cell adhesion molecule 2 | 1.00E-03 | -1.80 |
| SNHG15** | small nucleolar RNA host gene 15 | 1.00E-03 | -1.91 |
| ID3 | inhibitor of DNA binding 3, HLH protein | 1.00E-03 | -2.53 |

\* Genes with two UniProt reviewed annotations: both included
\*\* Genes with no UniProt reviewed annotation
Differential gene expression for 66 genes at $p < 0.001$ in SLE-P monocytes (as in Table 3).

**Figure 4.**
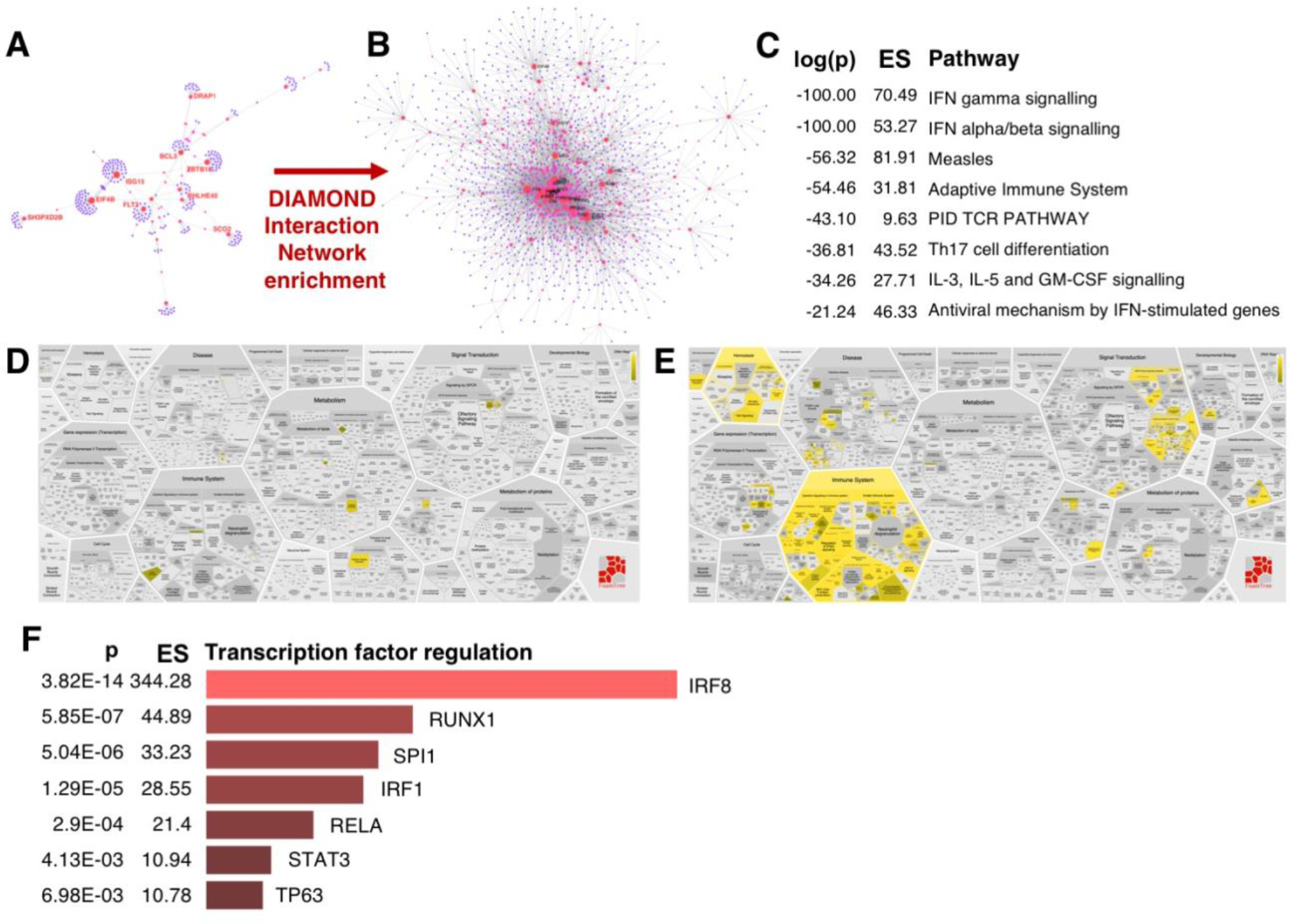
Enhanced interaction network analysis identifies pathways involved in adaptive immune system. **A**. Protein-protein interaction (PPI) network (Network Analyst, STRINGdb) of DE genes (≥1.5 fold change, p ≤ 0.001, n DE genes=66) analysed by RNA-seq as shown in Fig.1B. **B**. PPI network of DIAMOnD-extended DE list (n=268). **C**. Pathway enrichment analysis (Metascape) of genes in B. **D**. ReacFoam (Reactome) visualization of pathway enrichment analysis using Reactome curation plotted against biological process using genes in A. Yellow indicates enriched pathways. **E**. Pathway enrichment analysis as in D using DIAMOnD-extended gene list sown in B. **F**. Enrichment analysis (EnrichR: ChEA and ENCODE) of transcription factors associated with DIAMOnD-extended DE genes shown in B.

An enhanced PPI network analysis complements differential expression analyses by extending the original (seed) gene lists to include interacting protein neighbours within complexes or modules within the PPI-network that may be associated with a particular biological function (Ghiassian et al., 2015). In order to further identify *in silico* relevant functional pathways based on reported and predicted interactions iteratively added to the subnetwork, we next conducted an enhanced network analysis using the differentially expressed genes in SLE-P versus SLE-NP groups (Fig. 1B,C) as the seed set using the DIAMOnD algorithm (Ghiassian et al., 2015) (Fig. 4B).This curated a new gene list through the addition of proteins that are likely to be interacting with each seed gene at a high confidence level. As a result, a greater number of nodes (n=1407) and edges (n=3406) were predicted on the enhanced gene list (85 genes were excluded due to not being present in STRINGdb PPI-network reference database at an inclusion threshold of >0.8 confidence), visualised in Figure 4B.

Pathway enrichment analyses on the extended gene list (n=268) revealed, in addition to other pathways observed in the seed list (Fig. 1D), involvement of the adaptive immune system and T-cell receptor signalling (Fig. 4C,E; Table 8). Interestingly, recent studies identified T-cells as one of the dominant immune cell subsets in symptomatic human atherosclerotic plaques and described the transcriptional dysregulation in these cells (Fernandez et al., 2019). Our ongoing studies in a different cohort of juvenile onset-SLE (JSLE) patients has confirmed T-cells in JSLE patients show distinct global gene expression profiles, enriched in genes associated with apoptosis, T-cell activation and interferon signalling (Robinson et al., 2019). However, given known sex-specific differences in immune-cells (Márquez et al., 2020), it remains unknown whether the molecular identities of immune-cell subsets in male atherosclerotic plaques (Fernandez et al., 2019) are also relevant in women.

**Table 8:**
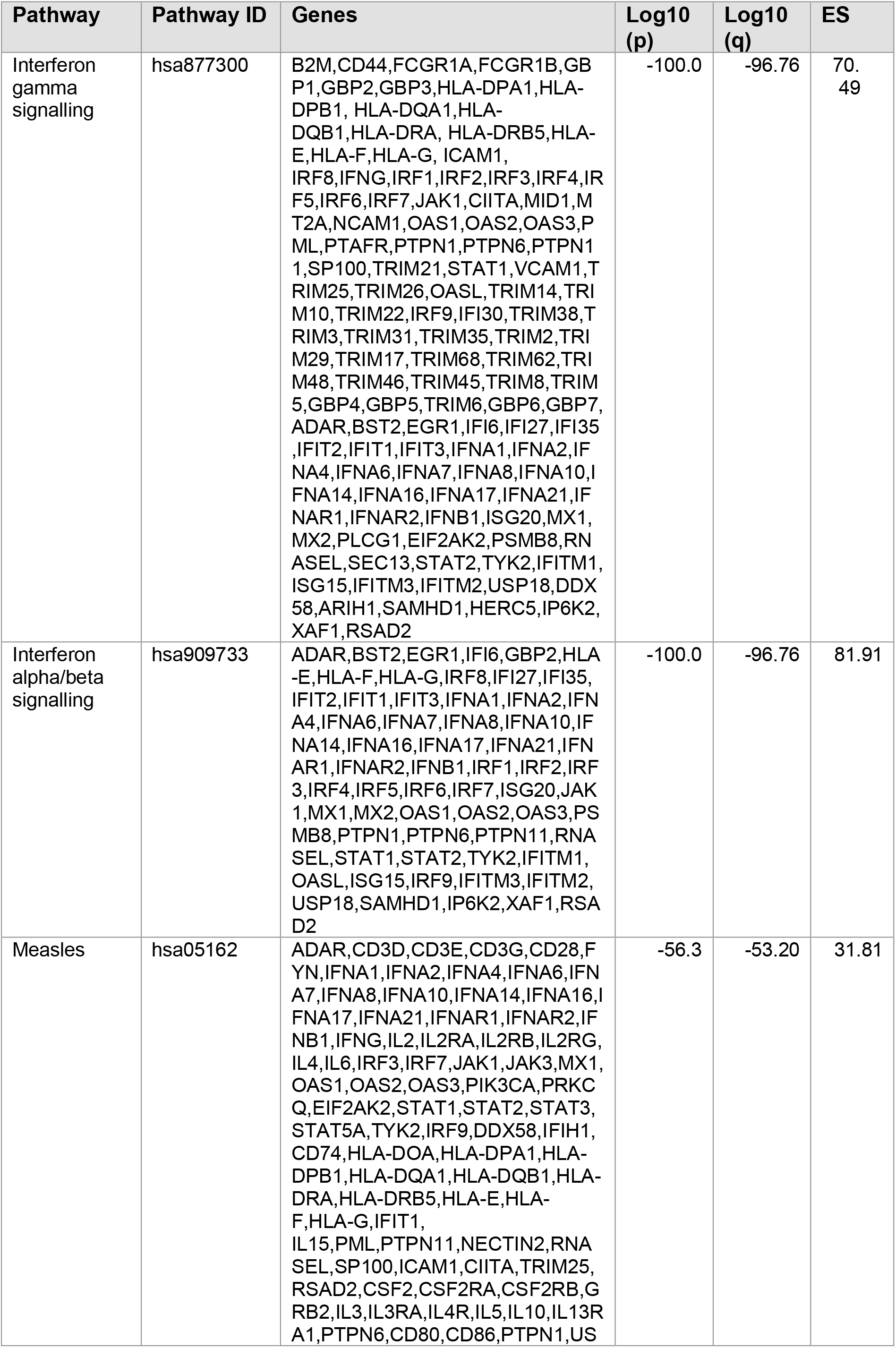
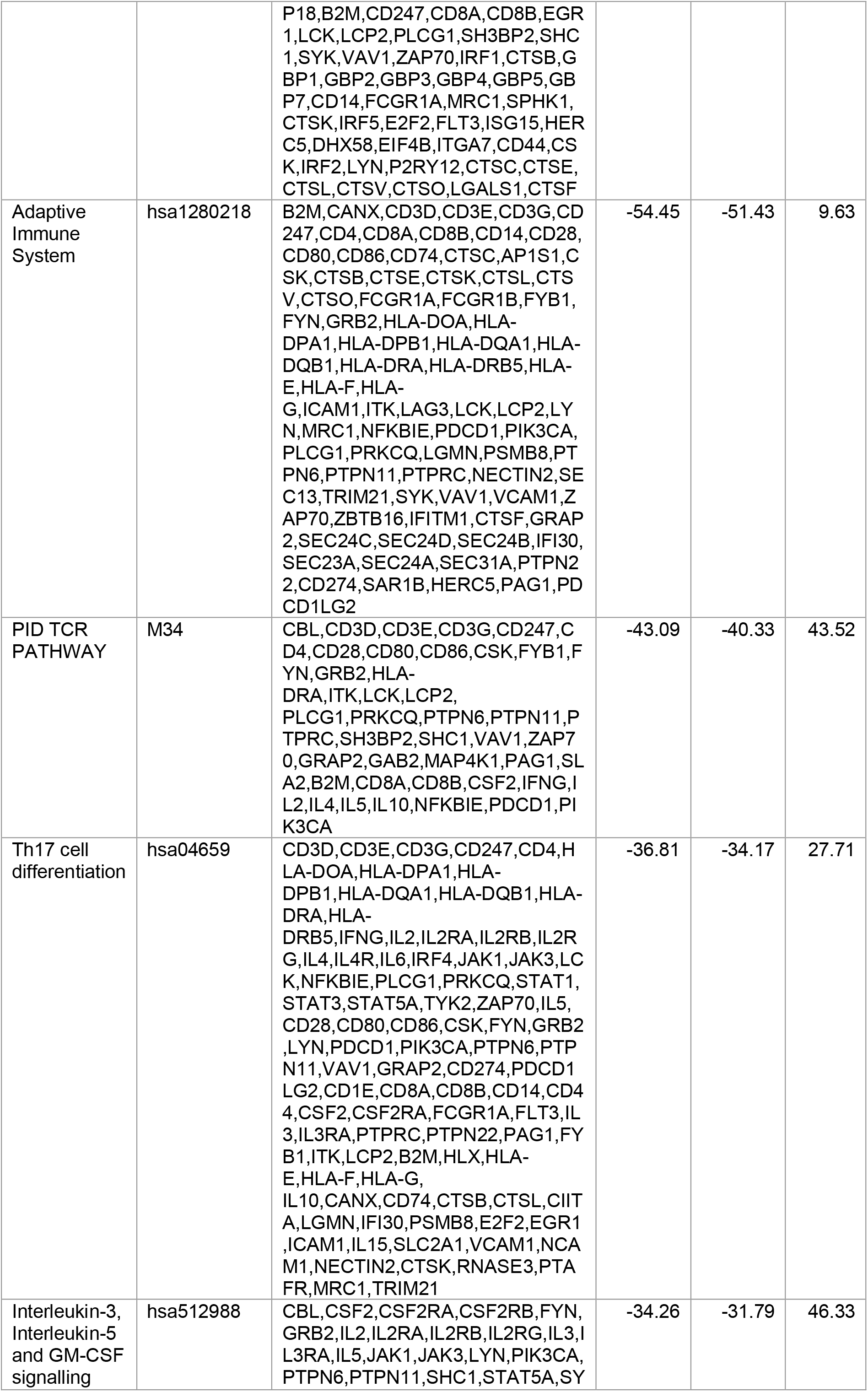
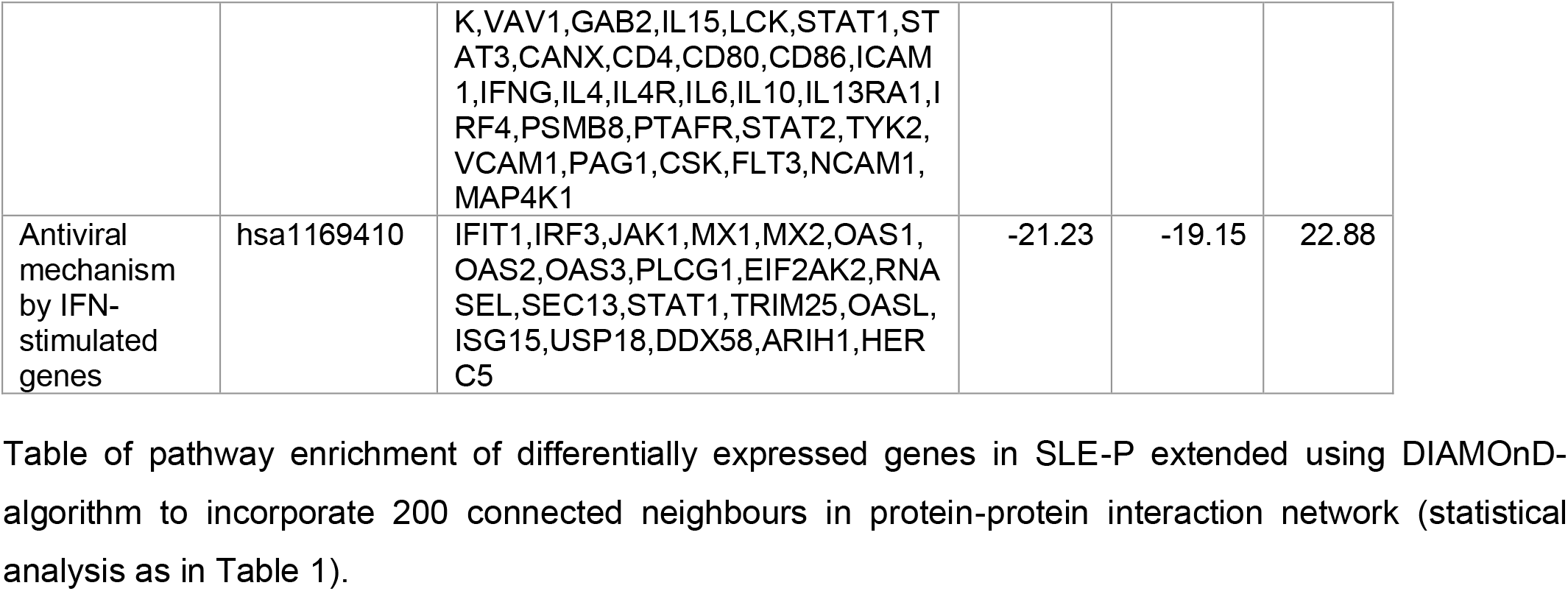
Metascape analysis of enriched Reactome/KEGG/Canonical/Corum pathways in DIAMOnD-extended differential gene expression list.

Transcription factor analysis with EnrichR revealed a significant regulator of the expanded gene set is IRF8 (Fig. 4F; Table 9). This transcription factor is required for the development and maturation of myeloid cells (monocytes, macrophages and dendritic cells), has shown anti-microbial functions and is implicated in cell activation in response to pro-inflammatory stimuli, including IFN (Jefferies, 2019; Salem et al., 2020). Notably, our work and that of others has implicated IRF8 in atherosclerosis plaque progression (Clément et al., 2018; Döring et al., 2012; Louie et al., 2019) with pro-or anti-atherogenic actions depending on the myeloid compartment it acts on or is signalled from. In addition, IRF8 variants have been associated with circulating monocyte counts (Crosslin et al., 2013), known to be associated with increased CVD risk (Coller, 2005) and other IRF variants have been linked to the presence of atherosclerotic plaques in lupus patients (Leonard et al., 2013).

**Table 9:** EnrichR analysis of transcription factors (TFs) regulating DIAMOnD-extended differential gene expression list.

| TF | Panel | Genes | p-value | Adj. p-value | Combined Score |
| --- | --- | --- | --- | --- | --- |
| IRF8 | ChEA | CD274,CD74,CIITA,DDX58,MX2,STAT2,IFI35,HLA-G,USP18,PSMB8,ISG20,IFIH1,HLA-DRA,TRIM26,IRF5,GBP2,TRIM21,IRF9 | 3.82E-14 | 3.97E-12 | 344.283 |
| RUNX1 | ChEA | SLC2A1,CSF2RB,PTPN22,ADARB1,SLA2,CD3E,SAMHD1,CD3D,CSF2RA,SLC2A6,HERC5,TRIM8,LGALS1,CTSL,ATG101,SH3BP2,E2F2,FFAR2,FYN,CTSE,SEC31A,STAT5A,IL10,IFNAR2,EGR1,SEC24B,STAT2,STAT3,VAV1,TRIM17,BHLHE40,BCL3,CD28,ID3,GRB2,LCP2,PDCD1,IRF9,LAT,LGMN | 5.85E-07 | 3.04E-05 | 33.232 |
| SPI1 | ChEA | SP100,CD82,PTAFR,CSF2RB,ICAM1,TRIM8,ADORA3,TRIM25,PIM3,CD14,FCGR1A,B2M,JAK1,LYN,PTPN1,SYK,MX2,STAT3,PML,VAV1,PSMB8,ISG20,BHLHE40,CANX,IRF2,ID3,PTPN6,IRF5,LCP2,CD68,NFKBIE,SEC24C,IFNAR1 | 5.04E-06 | 1.75E-04 | 28.555 |
| IRF1 | ENCODE | STAT5A,STAT1,SHC1,STAT2,MX1,IFI6,ISG15,IFI35,PSMB8,MT2A,IRF2,TRIM38,B2M,TRIM21 | 1.29E-05 | 3.36E-04 | 44.886 |
| RELA | ENCODE | CD74,CIITA,STAT1,TYK2,IL2RG,PSMB8,ICAM1,BST2,IFIH1,IRF1,BHLHE40,IRF2,IGFLR1,NFKBIE,B2M,JAK3,HLA-DQA1 | 2.93E-04 | 6.09E-03 | 21.405 |
| STAT3 | ENCODE | EGR1,RNASEL,WBP1,STAT1,SHC1,STAT2,STAT3,PDCD1LG2,PSMB8,HLA-E,ICAM1,HERC5,BST2,ISG20,MT2A,IRF1,BHLHE40,IRF2,IRF9 | 4.13E-03 | 7.16E-02 | 10.776 |
| TP63 | ChEA | CD82,PTAFR,ADARB1,SLA2,TRIM29,CTSK,GRAP2,SH3PXD2B,PLCG1,IFNAR2,EGR1,IL4R,SYK,STAT1,GAB2,PDCD1LG2,ALOX15B,PML,TPST1,ZAP70,IFNG,OAS2,IRF1,OAS3,IL2RB,PRKCQ,GRB2,IRF5,IRF6,CD44 | 6.98E-03 | 1.04E-01 | 7.975 |

Collectively, these molecular insights may contribute to guide future therapies to treat atherosclerosis in patients with SLE.

## Materials & Methods

### Patient cohort

Peripheral blood samples were collected from adult SLE (n=24) patients meeting the American College of Rheumatology (ACR) classification for SLE (Hochberg, 1997) or the Systemic Lupus International Collaborating Clinics (SLICC) criteria (Petri et al., 2012), attending a young adult rheumatology clinic at University College London Hospital (UCLH) and who had no previous history of CVD. Written informed consent was obtained and all patient identifiable data was removed; subsequent patient information was anonymised in accordance with relevant data protection legislation, including the EU General Data Protection Regulation (https://www.eugdpr.org/) and UK Data Protection Bill, 2018. This study was approved by the London - City & East Research Ethics Committee of the NHS 15-LO-2065.

### Vascular ultrasound scans

Carotid and femoral ultrasound scans were conducted to assess the presence of atherosclerotic plaques. Detailed patient characteristics and information regarding scanning can be found in Smith et al., (2016). Briefly, vascular ultrasound scans of the common carotid artery, carotid bulb, carotid bifurcation, common femoral artery and femoral bifurcation were performed bilaterally using the Philips IU22 ultrasound computer and the L9-3 MHz probe. IMT measurements were performed using QLAB Advanced Quantification Software® version 7.1 (Philips Ultrasound, Bothell, USA) twice within the course of 5 years. Plaque was defined as “a *focal thickening >1.2 mm that encroaches into the arterial lumen as measured from the media-adventitia interface to the lumen interface”* (Griffin et al., 2002). Patients who had at least one region fulfilling this description were included in the group with plaque (SLE-P). Patients were split into groups: with plaque reported in at least one region in both scans (SLE-P, n=15) and those remaining plaque-free after the second scan (SLE-NP, n=9). Peripheral blood samples (serum) were collected at the time of the second scan. SLE disease activity was determined by the British Isles Lupus Assessment Group (BILAG) Index. Clinical, demographic, serological, therapy, and disease activity characteristics for SLE-P and SLE-NP patients are shown in Table S1.

### RNA extraction, RNA sequencing and differential gene expression analysis

CD14+ monocytes were isolated from peripheral blood mononuclear cells (PBMCs) by FACS-sorting. PBMCs were washed in MACS buffer (1X PBS (Sigma), 2% FBS (Labtech) and 1mM EDTA (Sigma)) and stained with Zombie NIR™ Fixable Viability Kit (Biolegend) in PBS followed by staining for 30 minutes with appropriate flurochrome-labelled antibodies (Biolegend), in MACS buffer. Samples were sorted in MACS buffer using BD FACSAria flow cytometry cell sorter into collection media (1xPBS, 20% FBS). Total RNA from was extracted with TRIzol Reagent (Invitrogen). Sample concentration and purity was determined using a NanoDropTM 1000 Spectrophotometer. RNA integrity was confirmed using Agilent’s 2200 Tapestation. UCL Genomics (London, UK) performed library preparation and sequencing.

#### Sequencing

Libraries multiplexed in the same run were pooled in equimolar quantities, calculated from Qubit and Bioanalyser fragment analysis. Samples were sequenced on the NextSeq 500 instrument (Illumina, San Diego, US) using with 43bp paired end run with an 8bp UMI read.

#### Sequencing data and differential gene expression analysis

Sequencing data were demultiplexed and converted to fastq files using Illumina’s bcl2fastq Conversion Software. Samples were grouped by plaque status. To establish differences in gene expression between groups, sequence reads were aligned using STAR to the human hg38 reference genome. Gene count abundance was quantified using Partek E/M Annotation Model with default settings in Partek’s RNA Flow software. Differential expression analysis was performed using DESeq2-3.5 in RNA Flow, via Wald hypothesis testing and parametric fit typing and using the Benjamin-Hochberg method for multiple testing correction (see Robinson et al., 2019). IFN signature gene lists were obtained from published reports (IFN-GSEA (Panwar et al., 2020), IFN-Vital (El-Sherbiny et al., 2018). Publicly available differential gene expression datasets from atherosclerotic plaque macrophages were also obtained (Chai et al., 2018; Fernandez et al., 2019).

#### Data visualisation

Differential gene expression lists were shown as a volcano plots using VolcaNoseR [https://huygens.science.uva.nl/VolcaNoseR/] according to differential gene expression inclusion criteria (p<0.01, FC +/-1.5). Heatmaps depicting normalised gene counts for gene sets of interests were created using Partek RNA Flow or ClustVis [https://biit.cs.ut.ee/clustvis/] (Metsalu & Vilo, 2015) using correlation or Euclidian distance and average linkage for hierarchical clustering. Principle component analyses were conducted using normalised gene counts with ClustVis, using a linear transformation to interpret multivariate data into components, with components reporting the greatest amount of variance explained first. Unit variance scaling was applied to the rows and singular value decomposition with imputation was used to calculate the principal components. Venn diagrams were generated with BioVenn [https://www.biovenn.nl/index.php] (Hulsen et al., 2008).

### Pathway and transcription factor enrichment analysis of differential gene expression

Pathway enrichment analysis was performed using the web-based portal Metascape [https://metascape.org/] (Y. Zhou et al., 2019) on the DEG lists using inclusion threshold (FC < −1.5 or >+1.5, p<0.01). Pathways were considered enriched according to a significance threshold of p<0.01, minimum overlap of 3 genes, and minimum enrichment score of 1.5 from KEGG, Reactome Gene Sets, Canonical pathways and Corum structural complex repositories. Transcription factor enrichment was performed using EnrichR [https://amp.pharm.mssm.edu/Enrichr/] (Kuleshov et al., 2016); ChEA and ENCODE repositories were used as reference. Multiple list enrichment (Metascape) was used to compare pathways enriched across multiple datasets. And common pathways and conserved genes were presented as a circos plot.

### Protein-protein Interaction (PPI) network analysis using DIAMOnD

#### STRING database (STRINGdb)

Human protein interaction data was sourced from STRINGdb (version ii.0) (Szklarczyk et al., 2018), which utilises a confidence score scheme indicating the estimated likelihood a given interaction is biologically plausible. This includes direct (or physical) interactions and functional associations and combines experimental protein interaction data as well as prediction of potential interactors using co-expression analyses, automated text-mining and inclusion of gene orthology annotation data. Here, a filter was applied to exclude interactions with a score < 800 to reduce the likelihood of including false positives and filter low-scoring edges that tend to be attributable to noise. The human protein network was constrained by this parameter to include 14712 nodes (proteins) and 728090 edges (interactions); edges were unweighted and undirected.

#### DIAMOnD gene set expansion

To explore disease associated genes by considering the neighbourhood of seed genes within the PPI network, the diffusion algorithm DIAMOnD (Ghiassian et al., 2015) was applied to a subset of genes differentially expressed between SLE-P and SLE-NP at a stringent threshold of p<0.001 (n=66 genes, with 68 UniProt reviewed annotations, 12 seeds were excluded, DIAMOnD considers connectivity patterns associated with the disease genes based on a “connectivity significance” scoring scheme, that captures distantly connected proteins, obtained from STRINGdb). Any seed gene not present in the PPI-network (STRINGdb) at a confidence score of greater than 0.8 was excluded from the analysis. The confidence score indicates the likelihood a given protein is connected with another in the network, and whether this connection is biologically plausible. The high inclusion threshold used serves as a noise filter between two proteins. From the seed list, known disease genes were considered, then nodes from highly connected neighbours within a disease module were iteratively added. The combination of known disease genes and genes identified using DIAMOnD formed the disease module. An additional 200 genes were iteratively added forming the DIAMOnD-extended list.

#### DIAMOnD expansion enrichment and network visualisation

Pathway enrichment (Metascape) and transcription factor regulation (EnrichR) analysis were repeated on the DIAMOnD-expanded gene set as above. ReacFoam plots of pathway enrichment [https://reactome.org/reacfoam/] (Jassal et al., 2020) were obtained using Reactome pathway browser [https://reactome.org/PathwayBrowser/] to visualise pathways from Reactome ontology. Network Analyst [https://www.networkanalyst.ca] (G. Zhou et al., 2019) was used to visualise gene expression data within the context of a “generic” PPI-network.

## Data Availability

Transcriptomic data will be made available in an open access repository upon acceptance for publication in a peer reviewed journal

## Acknowledgments

This work was supported by BBSRC London Interdisciplinary Biosciences PhD consortium (LIDo) to L.W., BRC-NIHR-Research Project-BRC531/III/IPT/101350 to I.P.-T. & E.C.J., British Heart Foundation PhD studentship (FS/13/59/30649) to K.E.W., Lupus UK, The Rosetrees Trust (M409) and Multiple Sclerosis Society Project grant (076) to E.C.J. and Rosetrees Trust grant to to A.R.

## Author Contributions

I.P.-T. and E.C.J designed experiments, E.C. performed cell isolation and RNA extractions. L.W performed most data analysis, visualisations and prepared first draft of figures. G.R., K.E.W, contributed with data analysis and figures, I.P.T performed initial RNA-seq bioinformatic analysis. P.A. contributed to network analysis. A.R. performed patient recruitment and assessment; L.W., C.O., E.C.J. and I.P.-T., interpreted the data, and wrote the manuscript. IPT and ECJ conceived the study, secured the funding and supervised all aspects of the work.

